# Using early data to estimate the actual infection fatality ratio from COVID-19 in France

**DOI:** 10.1101/2020.03.22.20040915

**Authors:** Lionel Roques, Etienne Klein, Julien Papaïx, Antoine Sar, Samuel Soubeyrand

## Abstract

The number of screening tests carried out in France and the methodology used to target the patients tested do not allow for a direct computation of the actual number of cases and the infection fatality ratio (IFR). The main objective of this work is to estimate the actual number of people infected with COVID-19 and to deduce the IFR during the observation window in France. We develop a ‘mechanistic-statistical’ approach coupling a SIR epidemiological model describing the unobserved epidemiological dynamics, a probabilistic model describing the data acquisition process and a statistical inference method. The actual number of infected cases in France is probably higher than the observations: we find here a factor ×8 (95%-CI: 5–12) which leads to an IFR in France of 0.5% (95%-CI: 0.3 – 0.8) based on hospital death counting data. Adjusting for the number of deaths in nursing homes, we obtain an IFR of 0.8% (95%-CI: 0.45 – 1.25). This IFR is consistent with previous findings in China (0.66%) and in the UK (0.9%) and lower than the value previously computed on the Diamond Princess cruise ship data (1.3%).

## 1 Introduction

The COVID-19 epidemic started in December 2019 in Hubei province, China. Since then, the disease has spread around the world reaching the pandemic stage, according to the WHO World Health Organization (2020), on March 11. The first cases were detected in France on January 24. The infection fatality ratio (IFR), defined as the number of deaths divided by the number of infected cases, is an important quantity that informs us on the expected number of casualties at the end of an epidemic, when a given proportion of the population has been infected. Although the data on the number of deaths from COVID-19 are probably accurate, the actual number of infected people in the population is not known. Thus, due to the relatively low number of screening tests that have been carried out in France (about 5 over 10,000 people in France to be compared with 50 over 10,000 in South Korea up to March 15, 2020; Sources: Santé Publique France and Korean Center for Disease Control) the direct computation of the IFR is not possible. Based on the PCR-confirmed cases in international residents repatriated from China on January 2020, Verity et al. (2020) obtained an estimate of the infection fatality ratio (IFR) of 0.66% in China, and, adjusting for non-uniform attack rates by age, an IFR of 0.9% was obtained in the UK Ferguson et al. (2020). Using data from the quarantined Diamond Princess cruise ship in Japan and correcting for delays between confirmation and death Russell et al. (2020) obtained an IFR of 1.3%.

Using the early data (up to March 17) available in France, our objectives are: (1) to compute the IFR in France, (2) to estimate the number of people infected with COVID-19 in France, (3) to compute a basic reproduction number *R*_0_.

## 2 Materials and Methods

### 2.1 Data

We obtained the number of positive cases and deaths in France, day by day from Johns Hopkins University Center for Systems Science and Engineering Dong et al. (2020). The data on the number of tests carried out was obtained from Santé Publique France Santé Pulique France (2020). As some data (positive cases, deaths, number of tests) are not fully reliable (example: 0 new cases detected in France on March 12, 2020), we smoothed the data with a moving average over 5 days. Official data on the number of deaths by COVID-19 in France only take into account hospitalised people. About 728,000 people in France live in nursing homes (EHPAD, source: DREES DREES (2020)). Recent data in the Grand Est region (source: Agence Régionale de Santé Grand Est Agence Régionale de Santé Grand Est), report a total of 570 deaths in these nursing homes, which have to be added to the official count (1015 deaths on March 31).

### 2.2 Mechanistic-statistical model

The mechanistic-statistical formalism, which is becoming standard in ecology Roques et al. (2011); Roques and Bonnefon (2016); Abboud et al. (2019) allows the analyst to couple a mechanistic model that describes a latent variable, here an ordinary differential equation model (ODE) of the SIR type, and uncertain, non-exhaustive data. To bridge the gap between the mechanistic model and the data, the approach uses a probabilistic model describing the data collection process. A statistical method is then used for the estimation of the parameters of the mechanistic model.

#### Mechanistic model

The dynamics of the epidemic are described by the following SIR compartmental model:

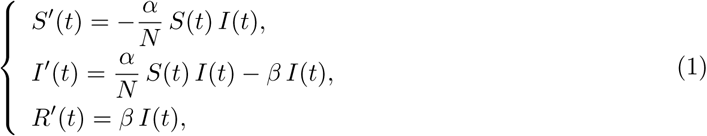

with *S* the susceptible population, I the infected population, *R* the recovered population (immune individuals) and *N* = *S* + *I* + *R* the total population, supposed to be constant. The parameter *α* is the infection rate (to be estimated) and 1/*β* is the mean time until an infected becomes recovered. Based on the results in Zhou et al. (2020), the median period of viral shedding is 20 days, but the infectiousness tends to decay before the end of this period: the results in He et al. (2020) show that infectiousness starts from 2.5 days before symptom onset and declines within 7 days of illness onset. Based on these observations we assume here that 1/*β* = 10 days.

The initial conditions are *S* (*t*_0_) = *N* − 1, *I* (*t*_0_) = 1 and *R*(*t*_0_) = 0, where *N* = 67 10^6^ corresponds to the population size. The SIR model is started at some time *t* = *t*_0_, which will be estimated and should approach the date of introduction of the virus in France (this point is shortly discussed at the end of this paper). The ODE system (1) is solved thanks to a standard numerical algorithm, using Matlab^®^ *ode45* solver.

Next we denote by *D*(*t*) the number of deaths due to the epidemic. Note that the impact of the compartment *D*(*t*) on the dynamics of the SIR system and on the total population is neglected here. The dynamics of *D*(*t*) depends on *I*(*t*) trough the differential equation:

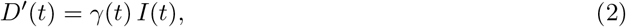

with *γ*(*t*) the mortality rate of the infecteds.

#### Observation model

We suppose that the number of cases tested positive on day *t*, denoted by 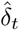, follow independent binomial laws, conditionally on the number of tests *n_t_* carried out on day *t*, and on *p_t_* the probability of being tested positive in this sample:

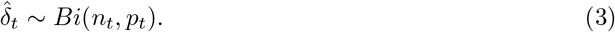

The tested population consists of a fraction of the infected cases and a fraction of the susceptibles: *n_t_* = *τ*_1_(*t*) *I*(*t*) + *τ*_2_(*t*) *S*(*t*). Thus,

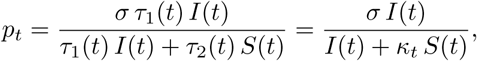

with *κ_t_*:= *τ*_2_ (*t*)/*τ*_1_ (*t*), the relative probability of undergoing a screening test for an individual of type *S vs* an individual of type *I* (probability of being tested conditionally on being *S* / probability of being tested conditionally on being *I*). We assume that the ratio *κ* does not depend on *t* at the beginning of the epidemic (i.e., over the period that we use to estimate the parameters of the model). The coefficient *σ* corresponds to the sensitivity of the test. In most cases, RT-PCR tests have been used and existing data indicate that the sensitivity of this test using pharyngeal and nasal swabs is about 63 – 72% Wang et al. (2020). We take here *σ* = 0.7 (70% sensitivity).

### 2.3 Statistical inference

The unknown parameters are *α*, *t*_0_ and *κ*. The parameter *γ*(*t*) is computed indirectly, using the estimated value of *I*(*t*), the data on *D*(*t*) (assumed to be exact) and the relationship (2). The likelihood 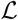 is defined as the probability of the observations (here, the increments 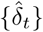) conditionally on the parameters. Using the observation model (3), and assuming that the increments 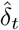 are independent conditionally on the underlying SIR process and that the number of tests *n_t_* is known, we get:

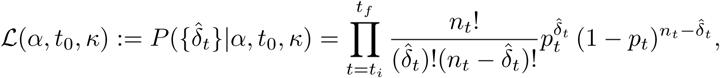

with *t_i_* the date of the first observation and *t_f_* the date of the last observation. In this expression 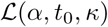 depends on *α*, *t*_0_, *κ* through *p_t_*.

The maximum likelihood estimator (MLE, i.e., the parameters that maximise 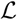), is computed using the BFGS constrained minimisation algorithm, applied to — 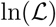, via the Matlab® function *fmincon.* In order to find a global maximum of 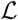, we apply this method starting from random initial values for *α*, *t*_0_, *κ* drawn uniformly in the following intervals: *α* ∈ (0,1), *t*_0_ ∈ (1, 31), (January 1st - 31th) and *κ* ∈ (0,1). The minimisation algorithm is applied to 10000 random initial values of the parameters.

The posterior distribution of the parameters (*α*, *t*_0_, *κ*) is computed with a Bayesian method, using uniform prior distributions in the intervals given above (a more informative prior distribution has also been tested, see Appendix). This posterior distribution corresponds to the distribution of the parameters conditionally on the observations:

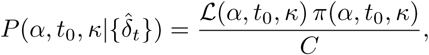

where *π*(*α,t*_0_,*κ*) corresponds to the prior distribution of the parameters (therefore uniform) and *C* is a normalisation constant independent of the parameters. The numerical computation of the posterior distribution is performed with a Metropolis-Hastings (MCMC) algorithm, using 4 independent chains, each of which with 10^6^ iterations, starting from random values close to the MLE (only the second half of the iterations are used to generate the posterior). The Matlab® codes are available as Supplementary Material.

The data 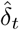 used to compute the MLE and the posterior distribution are those corresponding to the period from February 29 to March 17.

## 3 Results

### Model fit

To assess model fit, we compared the observations, i.e., the cumulated number of cases 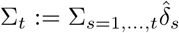, with the expectation of the observation model associated with the MLE (expectation of a binomial). Namely, we compared Σ*_t_* and 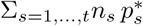 with

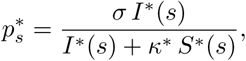

and *I*\*(*s*), *S*\*(*s*) the solutions of the system (1) (at time *s*) associated with the MLE. The results are presented in Fig. 1. We observe a good match with the data.

**Figure 1:**
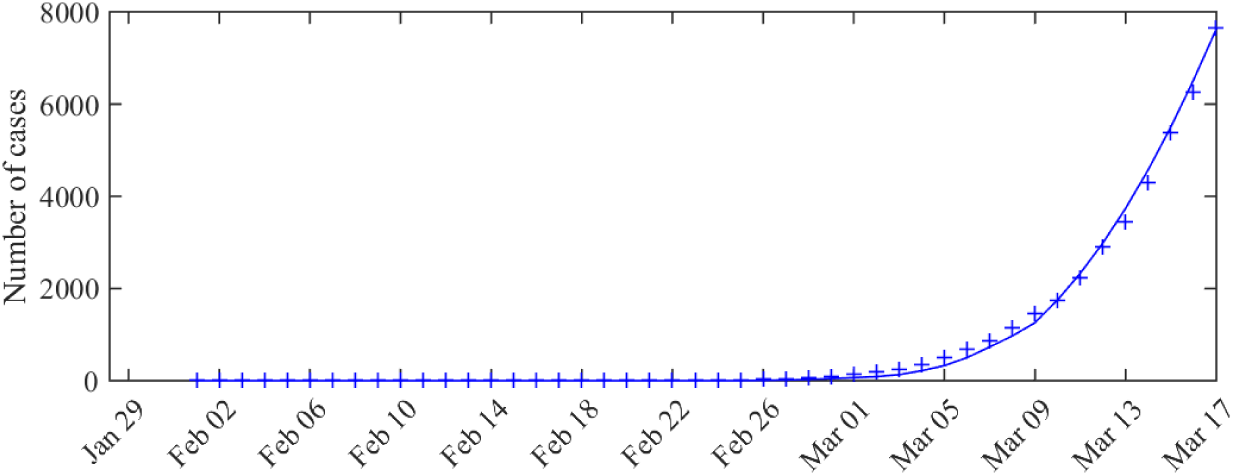
Expected number of observed cases associated with the MLE *vs* number of cases actually detected (total cases) The curve corresponds to cumulated values of the expected observation 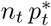 given by the model, and the crosses correspond to the data (cumulated values of 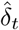).

### Infection fatality ratio and actual number of infected cases

Using the posterior distribution of the model parameters (the pairwise distributions are presented in Appendix, see Fig. A1), we computed the daily distribution of the actual number of infected peoples. Using the relation (2) together with the data on *D*(*t*) = Σ*_t_*, we deduce the distribution of the parameter *γ*(*t*), at each date. The IFR corresponds to the fraction of the infected who die, that is:

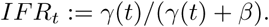

**Figure A1:**
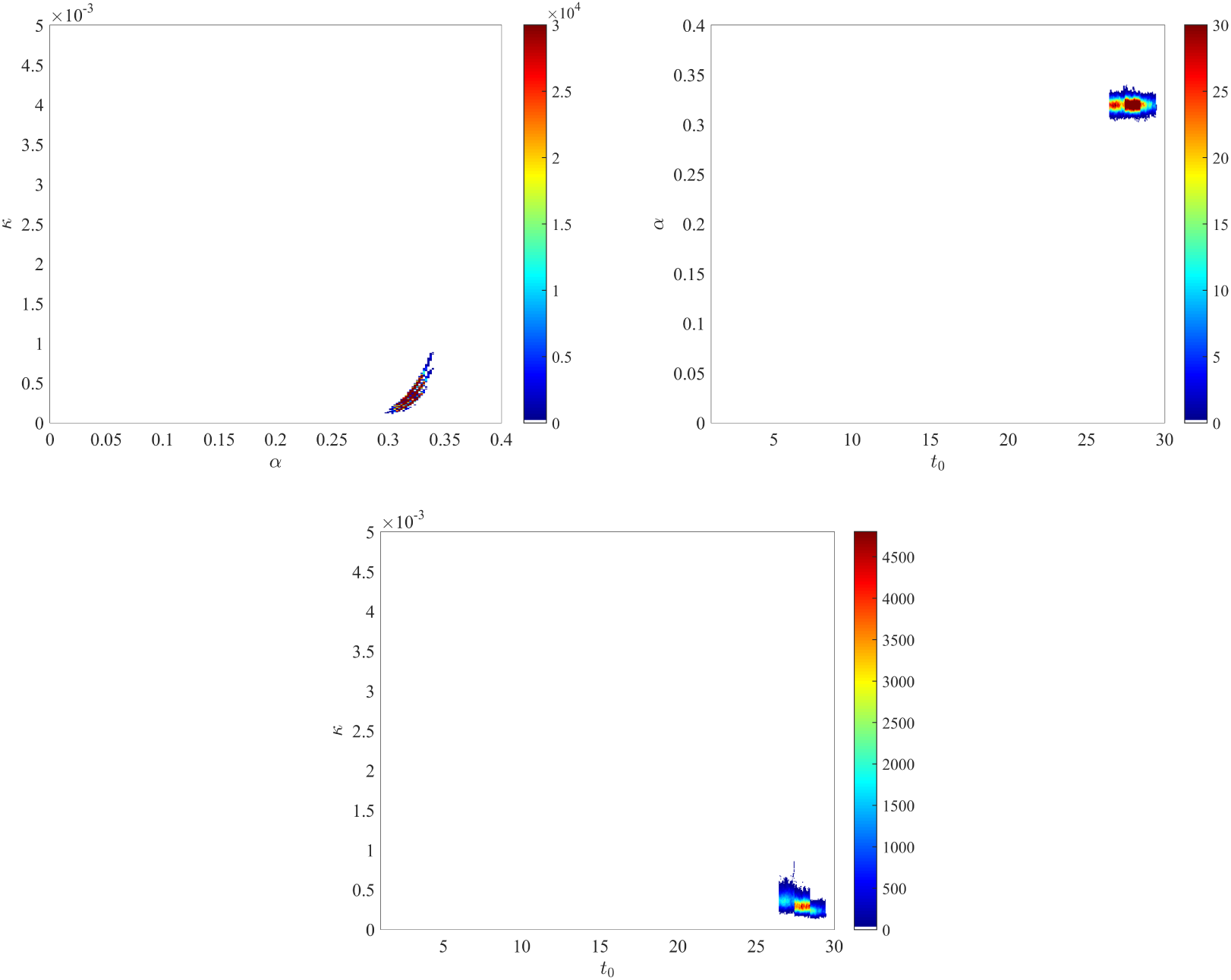
**Joint posterior distributions of** (*α*, *κ*), (*t*_0_, *α*) **and** (*t*_0_,*κ*).

**Figure A2:**
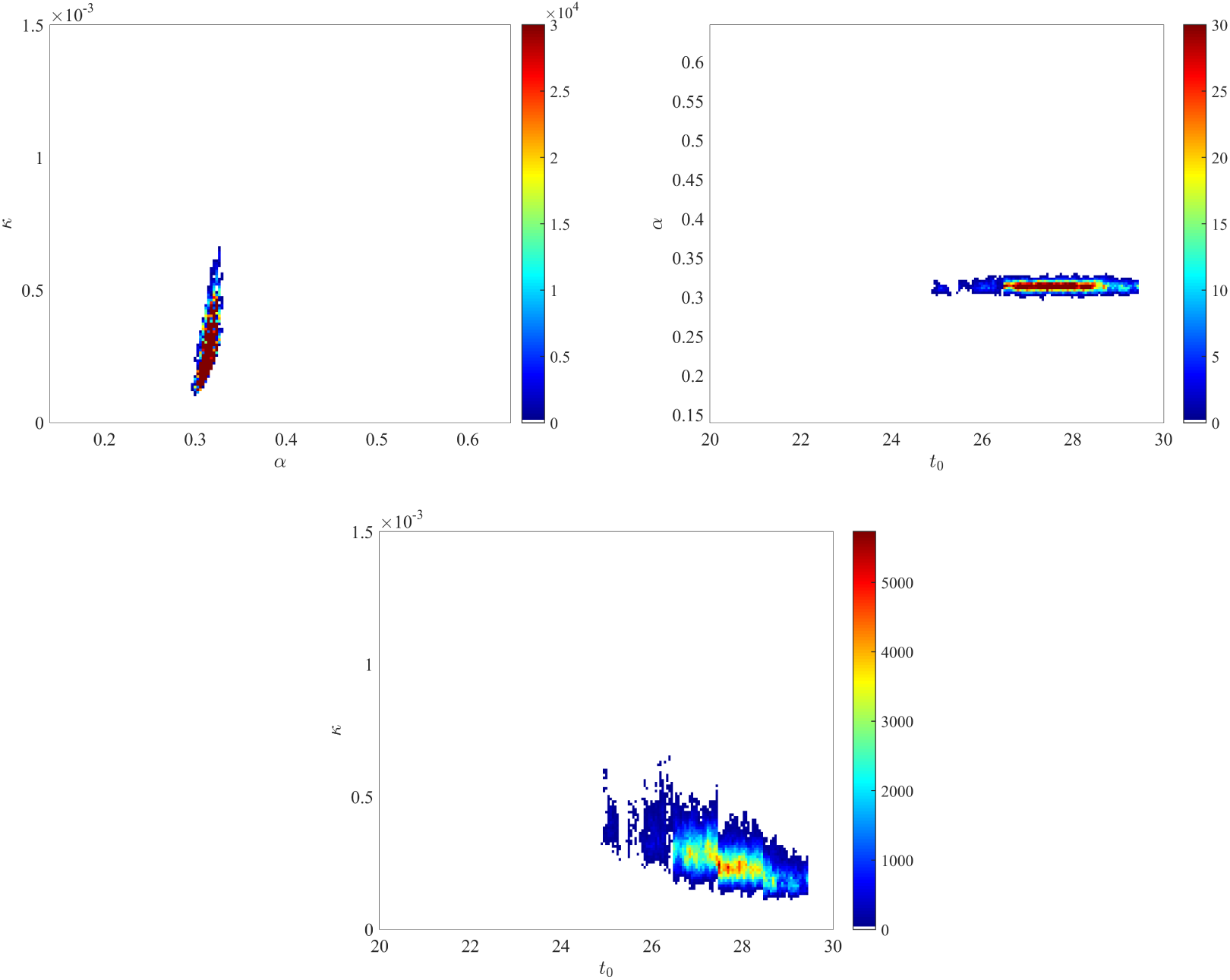
**Joint posterior distributions of** (*α*, *κ*), (*t*_0_, *α*) **and** (*t*_0_,*κ*) **obtained with a more informative prior**.

We thus obtain, on March 17 an IFR of 0.5% (95%-CI: 0.3 − 0.8), and the distribution of the IFR is relatively stable over time (see Fig. A3 in the Appendix).

**Figure A3:**
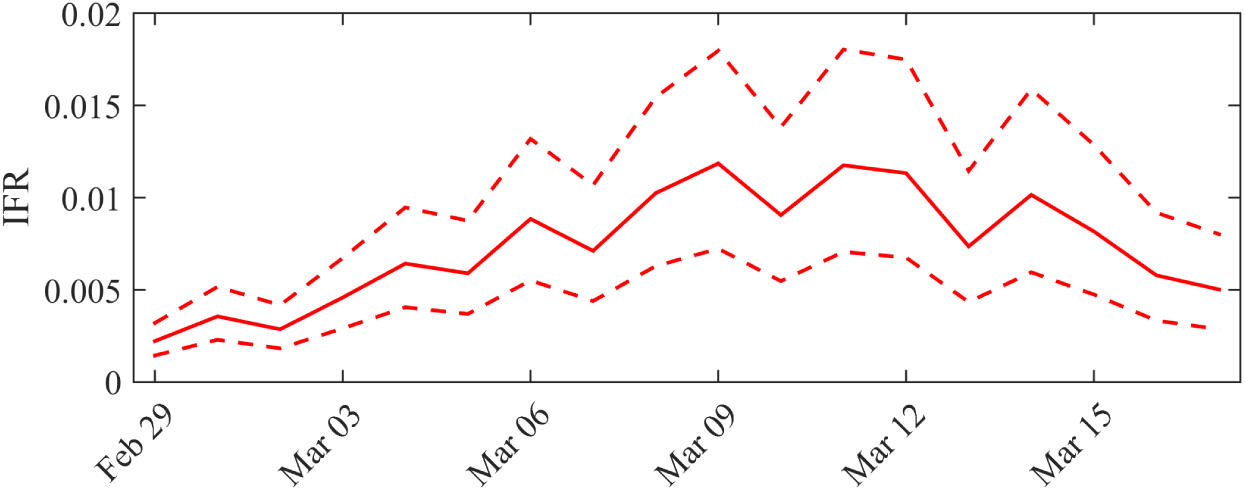
Dynamics of the IFR in France. Solid line: average value obtained from the posterior distribution of the parameters. Dotted curves: 0.025 and 0.975 pointwise quantiles.

Additionally, the distribution of the cumulated number of infected cases (*I*(*t*) + *R*(*t*)) across time is presented in Fig. 2. We observe that it is much higher than the total number of observed cases (compare with Fig. 1). The average estimated ratio between the actual number of individuals that have been infected and observed cases (*I*(*t*) + *R*(*t*))/Σ*_t_* is 8 (95%-CI: 5–12) over the considered period.

**Figure 2:**
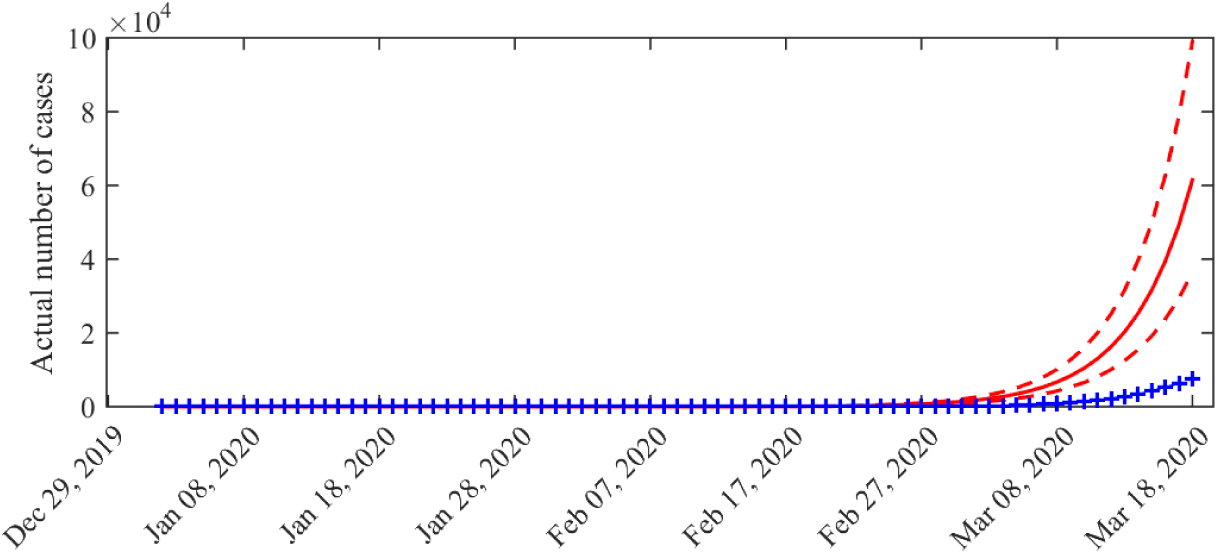
Distribution of the cumulated number of infected cases (*I*(*t*) + *R*(*t*)) across time. Solid line: average value obtained from the posterior distribution of the parameters. Dotted curves: 0.025 and 0.975 pointwise posterior quantiles. Blue crosses: data (cumulated values of 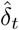).

### Taking into account the data in the nursing homes

The above computation of the IFR is based on the official counting of deaths by COVID-19 in France, which does not take into account the number of deaths in nursing homes. Based on the local data in Grand Est region, we infer that the IFR that we computed has been underestimated by a factor about (1015 + 570)/1015 ≈ 1.6, leading to an adjusted IFR of 0.8% (95%-CI: 0.45 − 1.25).

### Basic reproduction number

With SIR systems of the form (1), the basic reproduction number *R*_0_ can be computed directly, based on the formula *R*_0_ = *α*/*β* Murray (2002). When *R*_0_ < 1, the epidemic cannot spread in the population. When *R*_0_ > 1, the infected compartment *I* increases as long as *R*_0_ *S* > *N* = *S* + *I* + *R*. We computed the marginal posterior distribution of the basic reproduction number *R*_0_. This leads to a mean value of *R*_0_ of 3.2 (95%-CI: 3.1–3.3). The full distribution is available in the Appendix (Fig. A4).

**Figure A4:**
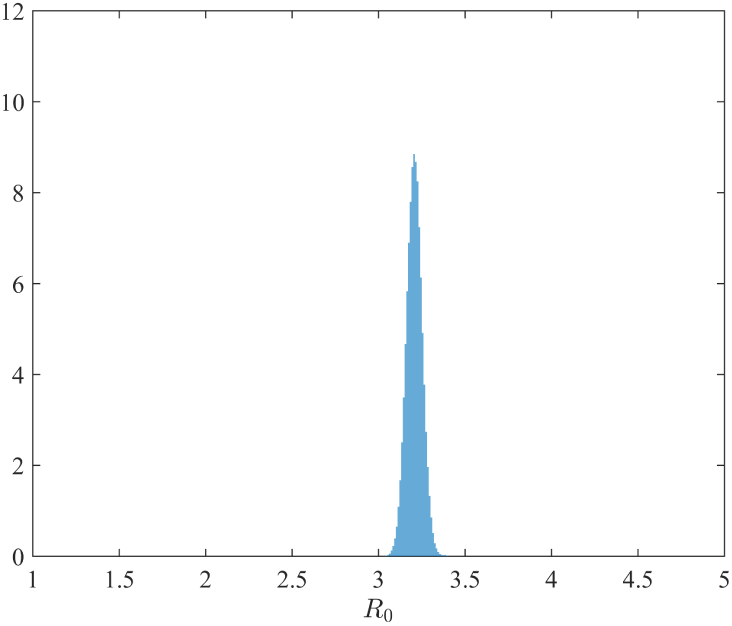
**Posterior distribution of the basic reproduction number** *R*_0_ **in France**.

### Sensitivity of the results with respect to the fixed model parameters

We computed the MLE with a larger infectious period (1/*β*) of 20 days estimated by Zhou et al. (2020). This leads to a much larger basic reproductive number *R*_0_ = 4.8 and a factor ×15 between the reported cases and the actual number of cases. However, the value of the IFR remains unchanged (0.5%). We also checked if the window width of the smoothing (moving average over 5 days) had an impact on our results. Computations of the MLE with a window width of 3 days (and *β* = 1/10) led to the same results as those presented above, namely a *R*_0_ = 3.2 and an IFR of 0.5%.

## 4 Discussion

### On the IFR and the number of infected cases

The actual number of infected individuals in France is probably much higher than the observations (we find here a factor ×8), which leads at a lower mortality rate than that calculated on the basis of the observed cases: we found here an IFR of 0.5% based on hospital death counting data, to be compared with a case fatality rate (CFR, number of deaths over number of diagnosed cases) of 2% on March 17. Adjusting for the number of deaths in the nursing homes, we obtained an IFR of 0.8%. These values for the IFR are consistent with the findings of Verity et al. (2020) (0.66% in China) and Ferguson et al. (2020) (0.9% in the UK). The value of 1.3% estimated on the Diamond Princess cruise ship Russell et al. (2020) falls above the top end of our 95% CI. This reflects the age distribution on the ship, which was skewed towards older individuals (mean age: 58 years), among whom the IFR is higher Ferguson et al. (2020); Russell et al. (2020).

The objective of our study was to estimate the IFR based on *early* data, before large scale surveys become available. By late April, new data and preliminary studies are available and can be compared to our results. An antibody study in New York released on April 24, 2020 shows about 14 percent tested positive, corresponding to 2.7 million cases, to be compared with the 271 000 confirmed cases and a statewide total of 15 500 deaths. This corresponds to an IFR of 0.6%. In France, another preliminary study conducted by Pasteur Institute Salje et al. (2020), and based on a joint estimate from French data up to April 14 (hospital death counting data) and from the Diamond Princess cruise ship data finds an IFR of 0.5% thus confirming our result. By April 28, the number of deaths from COVID-19 in Lombardy (Italy) is 13 575 (source: Ministero della Salute), for a population of 10 million people, showing that the IFR is *at least* of 0.14%. On the other hand, in South Korea where the number of detected cases rapidly reached a plateau, suggesting a small proportion of undetected cases, the ratio between the number of deaths and the number of positive cases is 244/10752 ≈ 2.3% (source: Johns Hopkins University Center for Systems Science and Engineering Dong et al. (2020)), which can be considered as an upper bound for the IFR, though overestimated.

If the virus led to contaminate 80% of the French population Ferguson et al. (2020), the total number of deaths to deplore in the absence of variation in the mortality rate (increase induced for example by the saturation of hospital structures, or decrease linked to better patient care) would be 336,000 (95%-CI: 192, 000 – 537,000), excluding the number of deaths in the nursing homes. This estimate could be corroborated or invalidated when 80% of the population will be infected, eventually over several years, assuming that an infected individual is definitively immunised. It has to be noted that measures of confinement or social distancing can decrease both the percentage of infected individuals in the population and the degree of saturation of hospital structures.

### *On the value of R*_0_

The estimated distribution in France is high compared to recent estimates (2.0-2.6, see Ferguson et al. (2020)) but consistent with the findings in Zhao et al. (2020) (2.24-3.58). A direct estimate, by a non-mechanistic method, of the parameters (*ρ*, *t*_0_) of a model of the form 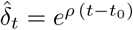 gives *t*_0_ = 36 (February 5) and *ρ* = 0.22. With the SIR model, *I′*(*t*) ≈ *I* (*α* − *β*) for small times (*S* ≈ *N*), which leads to a growth rate equal to *ρ* ≈ *α* − *β*, and a value of *α* ≈ 0.32, that is to say *R*_0_ = 3.2, which is consistent with our distribution of *R*_0_. Note that we have assumed here a infectiousness period of 10 days. A shorter period would lead to a lower value of *R*_0_.

### On the uncertainty linked to the data

The uncertainty on the actual number of infected and therefore the IFR are very high. We must therefore interpret with caution the inferences that can be made based on the data we currently have in France. In addition, we do not draw forecasts here: the future dynamics will be strongly influenced by the containment measures that will be taken and should be modelled accordingly.

### On the sensitivity of the results with respect to the fixed model parameters

We deliberately chose a parsimonious model with a few parameters to avoid identifiability issues. However, we needed to fix some parameter values. In particular, we assumed a mean duration of the infectious period (1/*β*) of 10 days. A much larger infectious period of 20 days (corresponding to the median period of viral shedding found in Zhou et al. (2020)) would lead to a much larger basic reproduction number *R*_0_ = 4.8 (but still within the range 1.4–6.49 described in Liu et al. (2020a)) and a factor ×15 between the reported cases and the actual number of cases. However, our main result on the IFR would remain unchanged (0.5%). We also assessed the sensitivity of the inference with respect to the prior knowledge, by proposing a set of more informative uniform prior distributions than the set specified in the main text. Overall, this prior modification does not significantly influence the posterior distributions; see Appendix A.

### On the hypotheses underlying the model

The data used here contain a limited amount of information, especially since the observation period considered is short and corresponds to the initial phase of the epidemic dynamics, which can be strongly influenced by discrete events. This limit led us to use a particularly parsimonious model in order to avoid problems of identifiability for the parameters. The assumptions underlying the model are therefore relatively simple and the results must be interpreted with regard to these assumptions. For instance, the date of the introduction *t*_0_ must be seen as an *efficient* date of introduction for a dynamics where a single introduction would be decisive for the outbreak and the other (anterior and posterior) introductions would have an insignificant effect on the dynamics.

A more complex epidemiological model of the COVID-19 epidemic in China has been proposed in Liu et al. (2020b), with an infectious class divided into several compartments (asymptomatic individuals, unobserved symptomatic infectious and observed symptomatic infectious). The authors use this model in Liu et al. (2020c) to make forecasts on the cumulative number of cases in China, while taking into account management strategies. In these two studies the authors emphasise the importance of being able to estimate the fraction of infectious cases that are not observed in order to forecast the dynamics of the epidemic. Our study, though based on a simpler SIR model, shows that this fraction can be estimated based on early data.

## Data Availability

All of the data in this manuscript are available from public sources: Johns Hopkins University Center for Systems Science and Engineering, Santé publique France, and Korean Center for Disease Control

https://github.com/CSSEGISandData/COVID-19

## Acknowledgements

This work was funded by INRAE: MEDIA Network.

## Author contributions statement

L.R., E.K.K., J.P., A.S. and S.S. conceived the model and designed the statistical analysis. L.R. and S.S. wrote the paper, L.R. carried out the numerical computations. All authors reviewed the manuscript.

## Abbreviations

IFR: Infection fatality ratio
CFR: Case fatality rate
ODE: Ordinary differential equation
ARS: Agence Régionale de Santé
WHO: World Health Organization
MLE: Maximum likelihood estimator
DREES: Direction de la recherche, des études, de l’évaluation et des statistiques

## Appendix

- The joint posterior distributions of the three pairs of parameters (*α*, *κ*), (*t*_0_, *α*) and (*t*_0_, *κ*) are depicted in Fig. A1.
- To check the robustness of our results with respect to the choice of the prior distribution, we also considered the case of a more informative prior. Namely, we assumed the following uniform prior distributions:

- *α* ∈ (0.14,0.65), corresponding to *β* × *R*_0_ with *β* = 1/10 and *R*_0_ values ranging between 1.4 and 6.49 (the range described in Liu et al. (2020a));
- *t*_0_ ∈ (20, 31) corresponding to an introduction during late January;
- *κ* ∈ (0,10^−2^), corresponding to a small probability of being tested for the susceptible cases, compared to the infected cases.

We obtained the posterior distributions shown in Fig. A2, based on two independent chains with 10^6^ iterations (only the second half of the iterations are used to generate the posterior). Overall, these distributions are relatively similar to those displayed on Fig. A1 and obtained with the prior distributions defined in the main text.
- The dynamics of the estimated distribution of the IFR are depicted in Fig. A3.
- The marginal posterior distribution of *R*_0_ is depicted in Fig. A4.

### Acknowledgements

We thank Laurent Desvillettes for his suggestion about the computation of a lower bound for the IFR based on data from the Lombardy province in Italy. We also thank the anonymous reviewers for their suggestions and comments.

## References

World Health Organization. WHO Director-General’s opening remarks at the media briefing on COVID-19 - 11 March *2020*. 2020.

R Verity, L C Okell, I Dorigatti, P Winskill, C Whittaker, N Imai, G Cuomo-Dannenburg, H Thompson, P Walker, H Fu, et al. Estimates of the severity of COVID-19 disease. medRxiv, 2020. doi: https://doi.org/10.1101/2020.03.09.20033357.

N M Ferguson, D Laydon, G Nedjati-Gilani, N Imai, K Ainslie, M Baguelin, S Bhatia, A Boonyasiri, Z Cucunuba, G Cuomo-Dannenburg, et al. Impact of non-pharmaceutical interventions (NPIs) to reduce COVID-19 mortality and healthcare demand. Imperial College, London, 2020. doi: https://doi.org/10.25561/77482.

T W Russell, J Hellewell, C I Jarvis, K van Zandvoort, S Abbott, R Ratnayake, S Flasche, R M Eggo, W J Edmunds, A J Kucharski, et al. Estimating the infection and case fatality ratio for coronavirus disease (COVID-19) using age-adjusted data from the outbreak on the Diamond Princess cruise ship, February 2020. Eurosurveillance, 25(12), 2020.

E Dong, H Du, and L Gardner. An interactive web-based dashboard to track COVID-19 in real time. The Lancet Infectious Diseases, 2020. doi: https://doi.org/10.1016/S1473-3099(20)30120-1. URL https://github.com/CSSEGISandData/COVID-19/.

Santé Pulique France. COVID-19: point épidémiologique du 24 mars 2020. 2020. URL https://geodes.santepubliquefrance.fr/.

DREES. 728 000 résidents en établissements d’hébergement pour personnes âgées en 2015. https://drees.solidarites-sante.gouv.fr/IMG/pdf/er1015.pdf, 2020.

Agence Régionale de Santé Grand Est. Dossier de presse - covid 19: point de situation dans le grand est, 2 avril 2020.

L Roques, S Soubeyrand, and J Rousselet. A statistical-reaction-diffusion approach for analyzing expansion processes. J Theor Biol, 274:43–51, 2011.

L Roques and O Bonnefon. Modelling population dynamics in realistic landscapes with linear elements: A mechanistic-statistical reaction-diffusion approach. PloS one, 11(3):e0151217, 2016.

C Abboud, O Bonnefon, E Parent, and S Soubeyrand. Dating and localizing an invasion from post-introduction data and a coupled reaction–diffusion–absorption model. Journal of Mathematical Biology, 79(2):765–789, 2019.

F Zhou, T Yu, R Du, G Fan, Y Liu, Z Liu, J Xiang, Y Wang, B Song, X Gu, et al. Clinical course and risk factors for mortality of adult inpatients with COVID-19 in Wuhan, China: a retrospective cohort study. The Lancet, 2020. doi: https://doi.org/10.1016/S0140-6736(20)30566-3.

X He, E HY Lau, P Wu, X Deng, J Wang, X Hao, Y Lau, J Y Wong, Y Guan, X Tan, et al. Temporal dynamics in viral shedding and transmissibility of COVID-19. medRxiv, 2020. doi: https://doi.org/10.1101/2020.03.15.20036707.

W Wang, Y Xu, R Gao, R Lu, K Han, G Wu, and W Tan. Detection of SARS-CoV-2 in different types of clinical specimens. Jama, 2020. doi: 10.1001/jama.2020.3786.

J D Murray. Mathematical Biology. Third edition, Interdisciplinary Applied Mathematics 17, Springer-Verlag, New York, 2002.

Henrik Salje, Cécile Tran Kiem, Noémie Lefrancq, Noémie Courtejoie, Paolo Bosetti, Juliette Paireau, Alessio Andronico, Nathanaël Hoze, Jehanne Richet, Claire-Lise Dubost, Yann Le Strat, Justin Lessler, Daniel Levy Bruhl, Arnaud Fontanet, Lulla Opatowski, Pierre-Yves Boëlle, and Simon Cauchemez. Estimating the burden of SARS-CoV-2 in France. April 2020. URL https://hal-pasteur.archives-ouvertes.fr/pasteur-02548181. preprint HAL-02548181.

S Zhao, Q Lin, J Ran, S S Musa, G Yang, W Wang, Y Lou, D Gao, L Yang, Daihai He, et al. Preliminary estimation of the basic reproduction number of novel coronavirus (2019-nCoV) in China, from 2019 to 2020: A data-driven analysis in the early phase of the outbreak. International Journal of Infectious Diseases, 92:214–217, 2020.

Ying Liu, Albert A Gayle, Annelies Wilder-Smith, and Joacim Rocklöv. The reproductive number of COVID-19 is higher compared to SARS coronavirus. Journal of travel medicine, 2020a. doi: https://doi.org/10.1093/jtm/taaa021.

Z Liu, P Magal, O Seydi, and G Webb. Understanding unreported cases in the 2019-nCov epidemic outbreak in Wuhan, China, and the importance of major public health interventions. MPDI Biology, 9(3):50, 2020b.

Z Liu, P Magal, O Seydi, and G Webb. Predicting the cumulative number of cases for the COVID-19 epidemic in China from early data. Mathematical Biosciences and Engineering, 2020c.

